# Forecasting Alzheimer’s Disease Progression with Deep Multimodal Learning: Integration of 3D MRI and Tabular Clinical Records via a Large Vision-Language Model

**DOI:** 10.64898/2026.01.06.26343479

**Authors:** Pengtao Dang, Fanyang Yu, Tingbo Guo, Jingwen Yan, Chi Zhang, Sha Cao, Somaye Hashemifar

## Abstract

**Background:** Accurate forecasting of Alzheimer’s Disease (AD) progression is critical for personalized patient management and clinical trial stratification. However, current predictive models often struggle to effectively integrate high-dimensional neuroimaging with longitudinal clinical data. We introduce AD-LLaVA-3D, a novel multimodal framework designed to bridge this gap by adapting large vision-language models for volumetric and temporal forecasting.

**Methods:** We leveraged the LLaVA-NeXT-Video architecture to treat 3D MRI volumes as temporal sequences, enabling the model to process volumetric imaging alongside longitudinal Tabular Clinical Records (TCR). The model was trained on the Alzheimer’s Disease Neuroimaging Initiative (ADNI) cohort (n=764) and evaluated using a rigorous patient-level split. We assessed its ability to forecast a suite of future clinical indicators (e.g., CDR-SB, MMSE) against traditional machine learning baselines (Lasso, Random Forest, Gradient Boosting) and specialized deep learning models (ResNet-3D, Med-Flamingo).

**Results:** AD-LLaVA-3D demonstrated superior predictive accuracy on the ADNI test set, achieving a Coefficient of Determination (*R*^2^) of 0.68 for the critical CDR-SB score, surpassing the best-performing baseline (*R*^2^ = 0.66). Crucially, in an independent external validation on the Open Access Series of Imaging Studies (OASIS) cohort (n=76), our model exhibited exceptional generalization (*R*^2^ = 0.82, *MSE* = 0.54), whereas comparison models showed significant performance degradation (*R*^2^ *<* 0.60).

**Conclusions:** This study presents the first application of a video-based multimodal architecture for AD progression forecasting. By effectively integrating 3D MRI with tabular clinical records, AD-LLaVA-3D offers a robust, generalizable tool for monitoring disease trajectories, significantly advancing predictive capabilities beyond current unimodal or static methods.

**Highlights:** First-in-Class Architecture: We introduce the first application of video-based Large Vision-Language Models (LVLMs) to interpret 3D volumetric MRI as a temporal sequence, capturing longitudinal neurodegeneration more effectively than static 3D-CNNs.

Robust External Validation: The model achieved superior predictive accuracy (*R*^2^ = 0.82) on an independent external cohort (OASIS), demonstrating exceptional generalization beyond the training population (ADNI).

Data-Efficient Multimodal Integration: We developed a novel prompting strategy that integrates sparse Tabular Clinical Records (TCR) without artificial imputation, allowing the model to leverage incomplete real-world medical history.

Clinical Trial Enrichment: By accurately forecasting future cognitive scores (CDR-SB, MMSE), AD-LLaVA-3D serves as a precise screening tool to identify ”rapid progressors” for clinical trials, potentially reducing failure rates in drug development.^1^

## 1 Introduction

Alzheimer’s Disease (AD) is a heterogeneous and progressive neurodegenerative disorder that presents a profound challenge to global healthcare systems(44; 34). The pathological complexity of AD, characterized by subtle metabolic changes, structural atrophy, and cognitive decline, necessitates a holistic diagnostic approach(15; 4; 46). Accurate forecasting of disease progression, predicting not merely the current state but the future clinical trajectory, is critical for stratifying patient risk and timing therapeutic interventions effectively. However, this predictive task is complicated by the multimodal nature of patient data, which spans high-dimensional volumetric imaging (MRI) and sparse, longitudinal Tabular Clinical Records (TCR)(14; 22; 38).

Historically, computational approaches to AD prognosis have been bifurcated. One stream of research focuses on Convolutional Neural Networks (CNNs), particularly 3D-CNNs like ResNet-3D, to extract spatial features from MRI scans (19; 16; 14; 49). While powerful at visual feature extraction, these models often function as ”black boxes” with limited interpretability and struggle to naturally integrate non-imaging modalities without complex, ad-hoc fusion layers. Conversely, traditional machine learning models (e.g., Lasso, Random Forest) excel at processing structured clinical data but fail to leverage the rich, spatially correlated information embedded in neuroimaging(47; 6; 48). This dichotomy limits the field’s ability to construct a unified patient representation that captures the interplay between neuroanatomical changes and clinical phenotype evolution.

The recent emergence of Large Vision-Language Models (LVLMs), such as LLaVA and GPT-4V, offers a promising avenue to bridge this gap (29; 42). By projecting visual and textual data into a shared semantic space, LVLMs demonstrate remarkable reasoning capabilities(13; 54; 57). However, the adaptation of these models to the medical domain remains nascent and faces two critical limitations. First, current medical LVLMs, such as Med-Flamingo (39), are predominantly designed for 2D static imaging (e.g., X-rays or single slices). They lack the native architectural capacity to process 3D volumetric data, which is essential for quantifying diffuse neurodegeneration in AD. Second, these models typically operate on static snapshots, ignoring the longitudinal temporal dimension required to model disease progression over time(27; 5).

To address these challenges, we introduce **AD-LLaVA-3D**, Fig 1 (c), a novel unified multimodal framework specifically engineered for the longitudinal forecasting of Alzheimer’s disease. Our core methodological insight is to adapt the architecture of video-based LVLMs (specifically LLaVA-NeXT-Video) to the domain of 3D medical imaging(31; 30; 29). By treating the slices of a 3D MRI volume as analogous to frames in a video sequence, our model effectively captures spatial continuity across the third dimension. Furthermore, unlike conventional classifiers, AD-LLaVA-3D functions as a generative forecaster: it takes a multimodal prompt, comprising the 3D MRI latent representation and a historical TCR trajectory, and generates a comprehensive text prediction of future clinical indicators (e.g., CDR-SB, MMSE) for a specified future time point.

**Figure 1:**
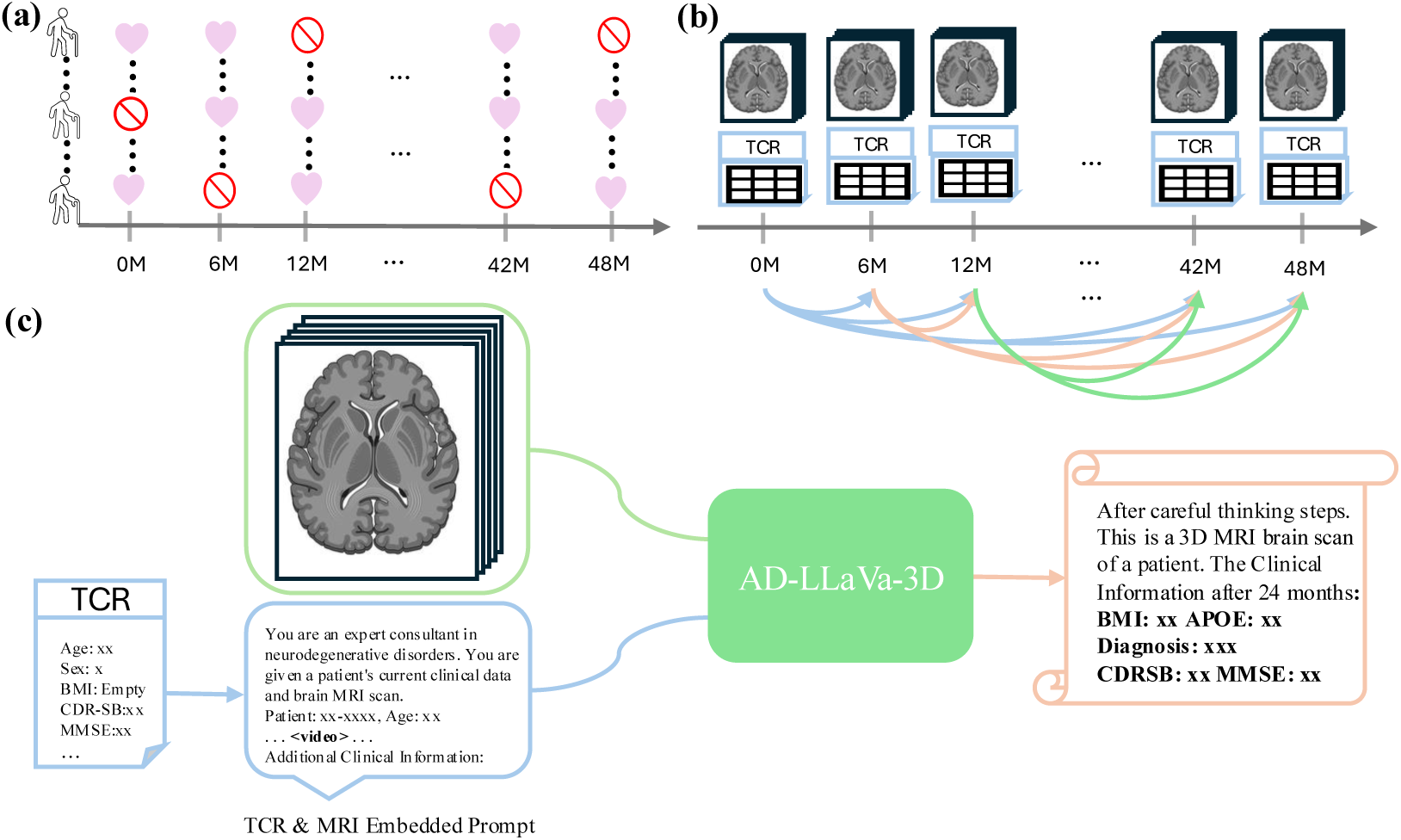
Overview of the AD-LLaVA-3D framework for longitudinal Alzheimer’s disease forecasting. (a) Challenge: Real-world clinical datasets often contain irregular or missing patient visits (indicated by the red symbols) across the longitudinal timeline (0–48 months). (b) Data Strategy: To address this, we construct valid predictive pairs by linking available historical data (MRI scans and Tabular Clinical Records [TCR]) to future time points. This allows the model to learn disease trajectories despite gaps in the timeline. (c) Multimodal Prediction: The AD-LLaVA-3D architecture functions as an ”AI Consultant.” It integrates a patient’s 3D MRI scan (processed as a volumetric sequence) with a structured text summary of their clinical history. The model then generates a comprehensive natural language report that forecasts the patient’s future diagnosis (e.g., MCI) and key cognitive scores (e.g., CDR-SB, MMSE).

This approach offers significant advantages in clinical realism. We address the pervasive issue of data sparsity in TCRs not through artificial imputation, but through a dynamic prompt design that omits missing fields, ensuring the model reasons only on high-fidelity, available data. We validate our framework through a rigorous experimental design using the Alzheimer’s Disease Neuroimaging Initiative (ADNI) dataset(23) for development and the Open Access Series of Imaging Studies (OASIS) dataset(33) for external validation.

The contributions of this work are summarized as follows:

1. **First Application of Video-LVLM Architecture for Volumetric AD Forecasting:** We propose a novel methodology that repurposes video-based large multimodal models to process 3D volumetric medical data. This allows AD-LLaVA-3D to capture complex neuroanatomical features without the information loss associated with 2D slice selection or the opacity of traditional 3D-CNNs. The code and checkpoints are available at github link https://github.com/ptdang1001/AD-LLaVa-3D
2. **Robust Integration of Sparse Clinical Modalities:** We develop a novel integration strategy for Tabular Clinical Records that bypasses the limitations of standard imputation techniques. By translating structured clinical data into natural language prompts, our model naturally handles the irregular and sparse nature of longitudinal medical history. This approach not only prevents the introduction of imputation noise but also significantly enhances data efficiency, enabling the model to learn complex disease trajectories from limited patient cohorts with superior generalization.
3. **Unified Longitudinal Prediction Framework:** Unlike baseline methods that require separate models for each clinical indicator, AD-LLaVA-3D serves as a unified foundation model. It simultaneously predicts a diverse suite of cognitive, functional, and behavioral metrics for future time points, effectively modeling the holistic trajectory of disease progression.
4. **Superior Generalization and Robustness:** Extensive empirical evaluation demonstrates that our approach significantly outperforms both unimodal baselines (ResNet-3D, Tabular ML) and state-of-the-art multimodal competitors (Med-Flamingo).

## 2 Related Work

### 2.1 Unimodal Approaches to Alzheimer’s Disease Prediction

#### Machine Learning on Tabular Clinical Records

The analysis of structured clinical data, encompassing demographics, genetic markers, and cognitive assessment scores, has long been a cornerstone of AD research. Traditional machine learning algorithms, particularly regression models like LASSO regression and ensemble methods such as Random Forest and Gradient Boosting, serve as the standard for prognostic modeling using this data modality (24; 43; 36; 26; 10). These models excel at identifying non-linear associations within structured features and are widely appreciated for their interpretability. However, their reliance on pre-defined, scalar feature vectors limits their capacity to ingest high-dimensional perceptual data. Consequently, they cannot account for the subtle, spatially distributed neuroanatomical changes, such as hippocampal atrophy or ventricular expansion, that often precede clinical symptoms (16; 41; 53). Furthermore, these models typically require complete data matrices, necessitating imputation strategies that can introduce noise when applied to sparse longitudinal records(45; 21; 3).

#### Deep Learning on Volumetric Neuroimaging

Parallel to tabular analysis, deep learning has revolutionized the interpretation of neuroimaging data. Convolutional Neural Networks (CNNs), specifically 3D architectures like ResNet-3D and DenseNet, have become the dominant paradigm for extracting spatial features from volumetric MRI (19; 52). Unlike slice-based 2D methods, 3D-CNNs preserve the spatial continuity of the brain structure, allowing for the detection of complex atrophy patterns. Despite their success in classification tasks (e.g., AD vs. CN), these models often struggle with longitudinal forecasting. They typically process a single time-point scan as a static input, lacking the inherent architectural mechanisms to model the temporal evolution of disease. Moreover, 3D-CNNs generally operate as ”black boxes,” providing limited transparency regarding the specific anatomical features driving a prognosis, which restricts their clinical adoption(1; 35; 25).

### 2.2 Multimodal Fusion Strategies

To bridge the gap between imaging and clinical data, recent studies have explored multimodal fusion techniques. The most common approach, often termed ”late fusion”, involves training separate unimodal networks (e.g., a CNN for MRI and an MLP for tabular data) and concatenating their output vectors before the final classification layer (12; 18; 56; 2). While this enables the simultaneous consideration of diverse data types, it fails to model the deep semantic interactions between modalities. For instance, a specific cognitive deficit recorded in the tabular data may directly correlate with a localized lesion in the MRI; late fusion methods often miss these fine-grained cross-modal dependencies(39; 28). Additionally, these architectures are rigid, requiring fixed-size inputs and struggling to adapt to the irregular, missing-data scenarios common in real-world clinical datasets (40).

### 2.3 Vision-Language Models in Medicine

The advent of Large Vision-Language Models (LVLMs) has introduced a new capability to process multimodal data in a unified semantic space. General-domain models have demonstrated the ability to reason over interleaved image and text inputs(54; 17; 9). In the medical domain, specialized adaptations such as LLaVA-Med (28) and Med-Flamingo (39) have shown promise in tasks like visual question answering (VQA) and report generation. These models leverage attention mechanisms to align visual features with textual clinical concepts dynamically.

However, a critical gap remains in the application of LVLMs to neurodegenerative disease forecasting. Existing medical LVLMs are predominantly designed for 2D static imaging (e.g., X-rays, pathology slides) and lack native support for the 3D volumetric data essential for neuroimaging. Current workarounds, such as selecting a single 2D slice, result in significant information loss. Furthermore, these models are typically trained for static analysis rather than longitudinal prediction(7; 20; 11). Our work addresses these limitations by adapting the architecture of video-based LVLMs (LLaVA-NeXT-Video) to the medical domain, effectively treating 3D MRI volumes as temporal sequences to capture both volumetric depth and longitudinal disease progression within a unified framework.

## 3 Method

### 3.1 Overview of the Model Architecture

AD-LLaVA-3D leverages the LLaVA-NeXT-Video architecture, a state-of-the-art large multimodal model, for the task of predicting Alzheimer’s disease (AD) progression, as shown in Fig 1 (C). The LLaVA framework connects a pre-trained CLIP ViT-L/14 vision encoder to a powerful Vicuna large language model (LLM) via a sophisticated multimodal projector (31; 8). This foundation is enhanced in LLaVA-NeXT-Video to process sequential visual data, making it uniquely suitable for our clinical application.

The model’s advanced video-handling capability is central to this research. Instead of adding specialized temporal modules, LLaVA-NeXT-Video processes a sequence of visual frames by concatenating their features into a single, long input for the LLM. The self-attention layers within the language model then perform a type of emergent hierarchical reasoning, identifying complex relationships and patterns across the entire sequence (31). We harness this functionality by treating a 3D MRI scan as a video-like data stream, where each consecutive 2D slice corresponds to a frame. This approach allows the model to analyze the full volumetric and anatomical context of the brain, interpreting the subtle, slice-to-slice structural changes that are critical hallmarks of AD.

### 3.2 Innovative Prompt Design for TCR Integration

A key innovation of this work is our prompting mechanism, which is designed not only to incorporate structured TCR data but also to elicit a form of clinical **chain-of-thought reasoning**(51; 32) from the model. We developed prompt formats that transform tabular health records into a coherent textual input. Crucially, the prompt strategically orders the predictive tasks: it first guides the model to forecast a group of related clinical indicators (such as the Age, Education, GLOBAL-CDR, and BMI). Only after establishing this broader clinical context does the prompt direct the model to predict the most critical and complex cognitive endpoints: the Clinical Dementia Rating Sum of Boxes (CDR-SB) and the Mini-Mental State Examination (MMSE). This step-by-step design encourages the model to build a more comprehensive understanding of the patient’s state before tackling the primary predictions(55; 37; 50), a process that proved highly effective in improving performance.

The fine-tuning process involved pairing these detailed, TCR-driven prompts with their corresponding 3D MRI scans. The model processes the MRI as a video-like sequence of slices, and this visual information is analyzed in conjunction with the clinical context provided by our text prompt. We then performed supervised fine-tuning on this combined dataset of AD patients, optimizing the end-to-end model to predict the full suite of future clinical scores. This integrated approach ensures that the model learns to establish direct correlations between MRI-derived brain anatomy and TCR-based health records, significantly enhancing its predictive power for AD progression and demonstrating a powerful application of a general-purpose multimodal model to a specific, complex clinical problem(5; 11; 7).

## 4 Experiments

This section details the comprehensive experimental framework designed to rigorously evaluate our AD-LLaVA-3D model for forecasting Alzheimer’s disease progression. We first describe the cohorts used: the primary ADNI dataset for training and internal validation, and the independent OASIS dataset for assessing the model’s external generalization capabilities. We then benchmark AD-LLaVA-3D’s performance against a diverse suite of baselines, including traditional machine learning models, a unimodal 3D-CNN, and a large vision-language model. Finally, we present in-depth ablation and sensitivity analyses to precisely quantify the synergistic contributions of fusing MRI and TCR data and to validate our architectural and data processing choices.

### 4.1 Dataset

This study utilizes data from two public databases: the Alzheimer’s Disease Neuroimaging Initiative (ADNI) and the Open Access Series of Imaging Studies (OASIS), as shown in Table 1. The primary cohort, sourced from ADNI, initially comprised 764 patients with longitudinal records containing both MRI scans and Tabular Clinical Record (TCR) data. For independent external validation to assess model generalizability, we used a cohort of 76 patients from the OASIS database. To leverage the temporal nature of the data for predictive modeling, we implemented a data augmentation strategy. For each patient, all possible forward-predicting pairs were generated from their sequential visits, where data from an earlier visit (*t*_*i*_) is used to predict outcomes at a later visit (*t*_*j*_ , where *j > i*), see Fig 1 (b). This process expanded the 764 ADNI patients into 6,326 predictive samples and the 76 OASIS patients into 642 samples. Each sample will have a unique anonymous patient name composed of random letters and numbers. The distinct template regions highlighted in yellow represent dynamic placeholders. For every sample, we fetch the corresponding patient data to populate these fields. Furthermore, we randomly sample one of four template architectures for each iteration; this variation prevents the model from overfitting to a specific syntax and enhances its ability to generalize across different input formats. Please forward to APPENDIX Fig S2, S3, S4, S5 for more prompt templates.

**Table 1:** Demographic, Clinical, and Longitudinal Characteristics of the Study Cohorts. Values are presented as Mean ± Standard Deviation (SD) for continuous variables and counts (percentage) for categorical variables. The ADNI cohort was partitioned at the patient level to prevent data leakage, while OASIS served as an independent external validation set. The “Clinical Progression” section highlights the dynamic nature of the cohort, distinguishing patients whose diagnosis remained stable from those who converted to a more severe stage during the study period.

| Characteristic | ADNI Cohort (Primary) |  |  |  | OASIS Cohort |
| --- | --- | --- | --- | --- | --- |
|  | Total<br>(n = 764) | Training<br>(n = 458) | Validation<br>(n = 152) | Testing<br>(n = 154) | External Val.<br>(n = 76) |
| **Demographics** |  |  |  |  |  |
| Age (years) | 75.3 ± 6.8 | 75.5 ± 6.4 | 74.5 ± 7.0 | 75.7 ± 7.6 | 69.8 ± 7.3 |
| Gender (Female, %) | 320 (41.9%) | 203 (44.3%) | 56 (36.8%) | 61 (39.6%) | 43 (56.6%) |
| Education (years) | 15.6 ± 3.0 | 15.6 ± 3.0 | 15.5 ± 3.1 | 16.2 ± 2.9 | 15.1 ± 3.1 |
| **Longitudinal Statistics** |  |  |  |  |  |
| Total Samples (Predictive Pairs) | 6,326 | 3,794 | 1,225 | 1,307 | 642 |
| Visits per Patient (Mean) | 4.4 ± 1.3 | 4.4 ± 1.3 | 4.3 ± 1.3 | 4.5 ± 1.2 | 4.1 ± 2.1 |
| Visits per Patient (Range) | 2 – 7 | 2 – 7 | 2 – 7 | 2 – 7 | 2 – 11 |
| Follow-up Duration (Months) | 28.1 ± 12.1 | 28.5 ± 12.5 | 27.5 ± 11.8 | 27.7 ± 11.2 | 61.8 ± 36.5 |
| **Clinical Status & Progression** |  |  |  |  |  |
| **Diagnosis at Baseline** |  |  |  |  |  |
| CN | 205(26.8%) | 129 (28.2%) | 43 (28.3%) | 33 (21.4%) | 60 (78.8%) |
| MCI | 471 (61.7%) | 271 (59.1%) | 91 (59.9%) | 109 (70.8%) | 13 (17.1%) |
| AD | 88 (11.5%) | 58 (12.7%) | 18 (11.8%) | 12 (7.8%) | 3 (4.1%) |
| **Clinical Progression** |  |  |  |  |  |
| Stable (No diagnosis change) | 475 (62.2%) | 290 (63.3%) | 98 (64.5%) | 87 (56.5%) | 58 (76.3%) |
| Converters (e.g., MCI → AD) | 289 (37.8%) | 168 (36.7%) | 54 (35.5%) | 67 (43.5%) | 18 (23.7%) |
| **Baseline Cognitive Scores** |  |  |  |  |  |
| CDR-SB | 1.7 ± 1.8 | 1.7 ± 1.9 | 1.7 ± 1.9 | 1.9 ± 1.7 | 0.6 ± 1.5 |
| MMSE | 26.8 ± 2.6 | 26.8 ± 2.7 | 26.9 ± 2.6 | 26.6 ± 2.5 | 28.7 ± 1.5 |

**Note:** CN: Cognitively Normal; MCI: Mild Cognitive Impairment; AD: Alzheimer’s Disease; CDR-SB: Clinical Dementia Rating Sum of Boxes; MMSE: Mini-Mental State Examination.

A rigorous preprocessing pipeline was established to manage data quality, starting with the significant sparsity in both datasets. We first performed an initial filtering step, removing patients with only one visit and excluding any individual visits that lacked a corresponding MRI scan. To handle missing values for certain key indicators, we then implemented a conservative filling strategy: a missing value at one visit was filled using the value from the nearest temporal visit, but only if that visit occurred within a 90-day window (either forward or backward). For all other fields, we deliberately avoided general imputation; instead, any field with a remaining missing value was simply omitted from that sample’s textual prompt. However, for the baseline models that require a complete numerical input, specifically the conventional regression models, the ResNet-3D model, and the vision-only ablation model, any remaining missing values were imputed with zero. After these cleaning steps, the ADNI dataset was partitioned at the patient level into training (60%), validation (20%), and testing (20%) sets, resulting in 3,794 training, 1,225 validation, and 1,307 testing samples. Each pair was treated as a distinct data point with a unique identifier to enhance generalization.

The model was trained and validated exclusively on the ADNI training and validation sets. Its performance was then assessed on two separate, entirely unseen datasets: first, the held-out ADNI test split, and second, the complete OASIS dataset, which functioned as an external validation cohort. Crucially, the model was never exposed to any data from the OASIS cohort during any phase of its development, ensuring a robust and unbiased evaluation of its ability to generalize to a different patient population and data distribution.

### 4.2 Evaluation Metrics and Predictive Targets

We assessed the model’s predictive accuracy using two standard regression metrics: the **Coefficient of Determination (R**^2^) and **Mean Squared Error (MSE)**. R^2^ quantifies the proportion of variance in the target outcomes that is explained by the model, providing a measure of goodness-of-fit where a higher value indicates better performance. MSE measures the average squared difference between the predicted and actual values, where a lower value signifies a more accurate model.

For each data pair, the model is trained to predict a diverse set of clinical outcomes for the future time point (*t* _*j*_ ). These targets include cognitive and functional assessments, such as the **Clinical Dementia Rating Sum of Boxes (CDR-SB)**, **Mini-Mental State Examination (MMSE)**, **Apolipoprotein E (APOE)**, **Global Clinical Dementia Rating Score (Global CDR)**; and general health metrics, including **Body Mass Index (BMI)**.

### 4.3 Baseline Models

To evaluate the effectiveness of AD-LLaVA-3D. The trained model was benchmarked against a diverse set of baseline models. We compared our results against traditional machine learning regression models that use tabular data, deep learning model that uses imaging data and large multi-modal model uses image and text data.

- **Classical Regression Models:** To establish strong baselines using only structured TCR data, we trained three classical regression models. These included **Lasso** regression, a linear model known for its feature selection capabilities, and two powerful ensemble learning techniques: **Random Forest (RF)**, a bagging ensemble method that builds numerous decision trees on different data subsets and averages their predictions to reduce variance, and **Gradient Boosting Decision Tree (GBDT)**, a state-of-the-art boosting ensemble method that constructs models sequentially to correct the errors of prior ones. These models were trained on the tabular TCR features from an initial time point to forecast clinical indicators at a future visit. A key characteristic of this approach is that it requires training a separate, independent model for each clinical indicator we aimed to predict.
- **ResNet-3D:** As a strong imaging-only baseline, we employed a pretrained **ResNet-18-3D** model (19). This architecture extends the highly successful 2D Residual Network (ResNet) framework to handle volumetric data by replacing 2D convolutions with 3D convolutions. This adaptation allows the model to learn hierarchical spatial features directly from the raw 3D MRI scans, making it a powerful standard for 3D computer vision tasks. In our implementation, the ResNet-3D network served as a feature encoder to generate a compact vector representation of each patient’s brain scan. Following this encoder, and similar to our traditional machine learning baselines, a separate regression head was attached and trained for each individual clinical indicator, treating each prediction as an independent task.
- **Med-Flamingo:** To provide a state-of-the-art benchmark for multimodal forecasting, we fine-tuned **Med-Flamingo**, a powerful, open-source vision-language model specifically adapted for the medical domain. Med-Flamingo is built upon the general-purpose Flamingo architecture, which connects a pre-trained vision encoder to a large language model using sophisticated gated cross-attention layers. This design allows the language model to fluidly incorporate visual information from one or more images when processing text. The model was further pre-trained on a massive corpus of paired medical images and texts, making it a highly relevant and challenging baseline. For our study, we fine-tuned this model on the same combined MRI and TCR-embeded prompt dataset to create a direct comparison, allowing us to fairly assess the performance of our specialized architecture against a leading approach in multimodal medical AI.

As Fig 2 shown, our AD-LLaVA-3D demonstrated superior performance across all evaluation metrics, consistently outperforming a range of baseline models in forecasting key clinical indicators. As detailed in Figure 4, for the critical CDR-SB score, AD-LLaVA-3D achieved an R^2^ of 0.68 and an MSE of 2.34. This marks a significant improvement over the state-of-the-art multimodal baseline, Med-Flamingo, which yielded an R^2^ of 0.46 and an MSE of 3.27. Notably, our model also surpassed powerful machine learning methods specifically designed for regression, including Gradient Boosting (R^2^ 0.66, MSE 2.45) and Random Forest (R^2^ 0.64, MSE 2.47), while the imaging-only ResNet-3D model proved to be less effective for this complex, multi-value prediction task. A key advantage of our approach is its architectural efficiency; AD-LLaVA-3D operates as a single, unified model that predicts all clinical indicators at once. This contrasts with the traditional machine learning baselines, which require training separate models for each individual indicator, and the ResNet-3D model, which necessitates multiple regression heads. The superior predictive accuracy, evident across the full cohort as well as in stratified analyses by gender and time intervals, underscores the efficacy of our unified multimodal approach for Alzheimer’s disease progression forecasting.

**Figure 2:**
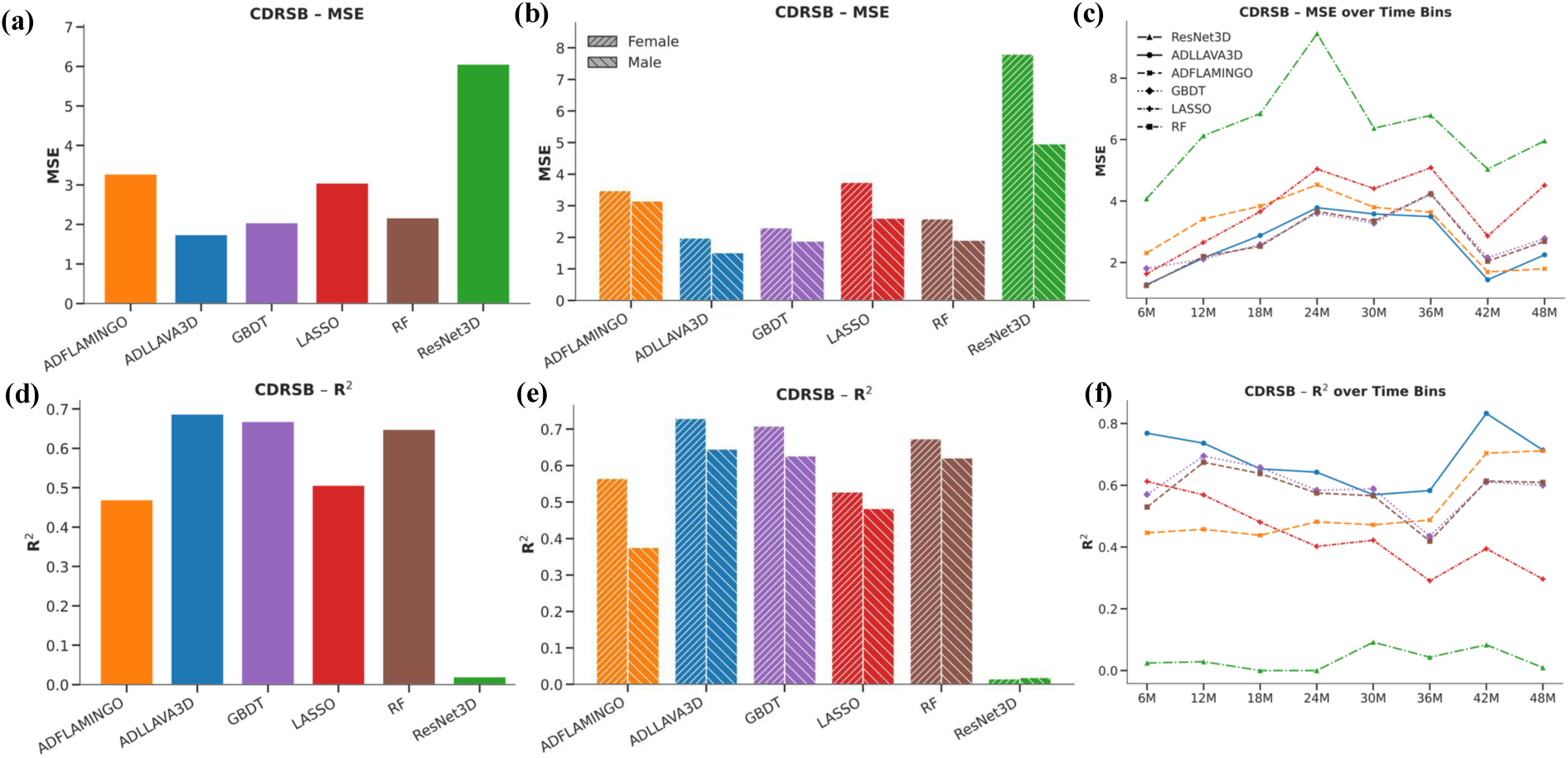
Performance evaluation of CDR-SB forecasting on the ADNI cohort, comparing overall predictive accuracy (MSE and *R*^2^), gender-specific robustness, and longitudinal stability across future time points against baseline models.

To rigorously evaluate the model’s generalization capabilities, we performed an external validation on the unseen OASIS dataset, Fig 3. This dataset was curated with a stricter filtering protocol where no missing indicator values were permitted, resulting in a higher-quality cohort compared to the ADNI training set which allowed for some data sparsity to maintain sample size. On this challenging, AD-LLaVA-3D demonstrated generalization ability. For the CDR-SB score, our model achieved an exceptional R^2^ of 0.82 and an MSE of 0.54. In stark contrast, the baseline models exhibited a significant performance degradation. The Med-Flamingo model’s performance collapsed, with its R^2^ dropping to 0.11 (MSE 2.69). Similarly, the conventional machine learning models showed poor generalizability; even the strongest among them, Random Forest, only reached an R^2^ of 0.60 (MSE 1.22), while Gradient Boosting and Lasso performed poorly with R^2^ values of 0.35 and 0.32, respectively. This substantial performance gap on the external validation set underscores the robustness and superior generalization capacity of our unified multimodal architecture.

**Figure 3:**
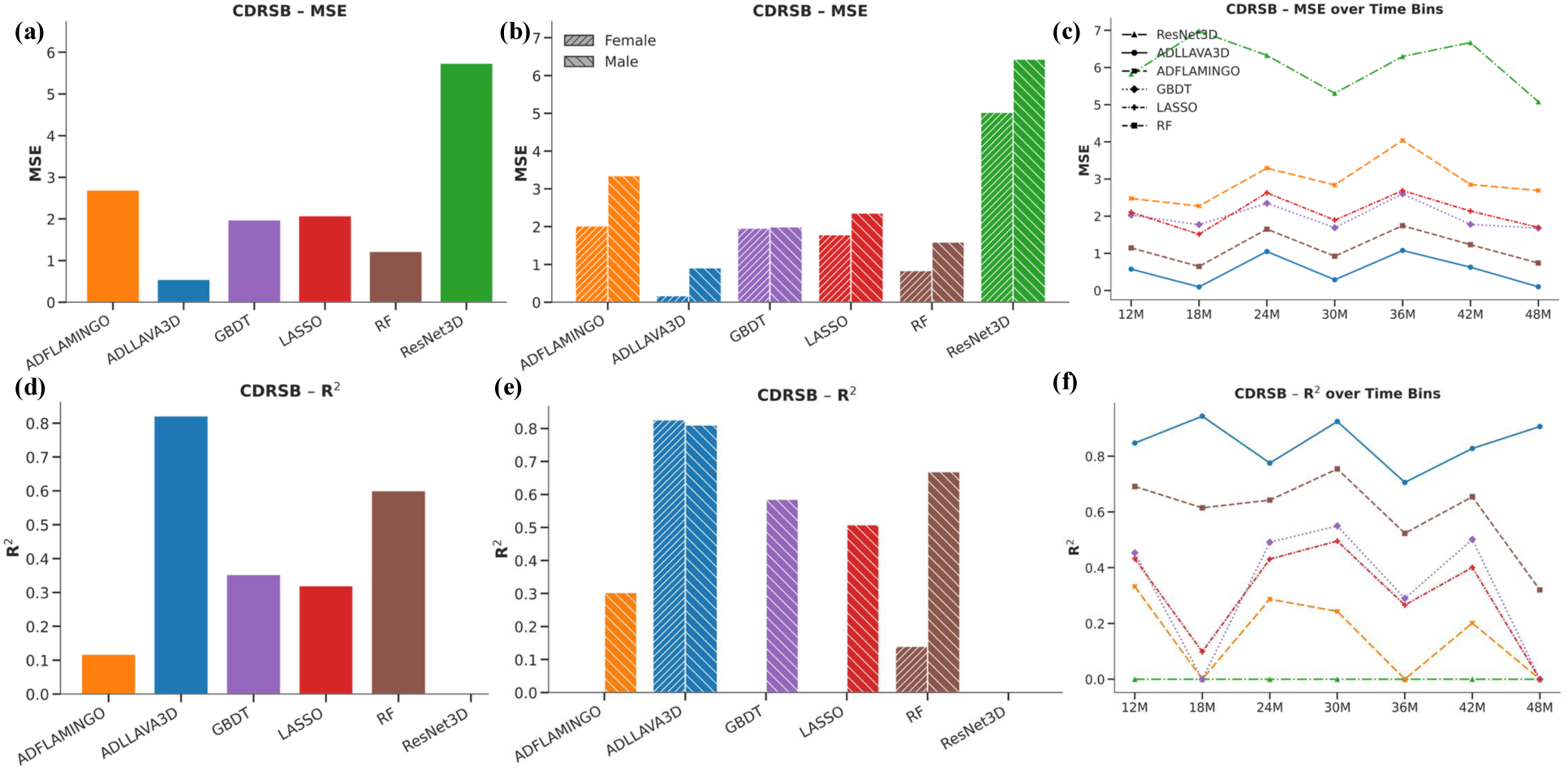
Performance evaluation of CDR-SB forecasting on the OASIS cohort, comparing overall predictive accuracy (MSE and *R*^2^), gender-specific robustness, and longitudinal stability across future time points against baseline models.

### 4.4 Ablation Studies

To rigorously analyze our approach, we conducted two sets of ablation studies. The first study quantifies the contribution of each data modality, while the second assesses the model’s sensitivity to different MRI slice sampling strategies.

#### 4.4.1 Ablation on Data Modalities

To isolate and quantify the impact of each data source, we conducted an ablation study by evaluating the following unimodal configurations:

- **Vision-Only:** This model isolates the visual pathway. The Vision encoder first processes the MRI data to generate a feature representation. This representation is then fed into multiple, separate neural network heads, with each head specifically trained to predict one of the clinical indicators (e.g., CDR-SB, MMSE), bypassing the language model entirely. As this architecture requires a complete numerical feature vector for the regression heads, any missing values in the corresponding TCR data were imputed with zero.
- **Languag e-Only:** In this configuration, only the Vicuna-7B large language model component was utilized. The model was provided with the detailed, TCR-driven text prompt but received no corresponding visual input from the MRI scan. This setup measures the predictive power derived exclusively from the structured and unstructured textual data, as processed by the LLM.

As Fig 4 (a) and (b) shown, we conducted an ablation study to dissect the individual contributions of the vision (MRI) and language (TCR) modalities within our framework. The results clearly demonstrate the synergistic effect of multimodal data fusion. On the ADNI test set, the vision-only model, consisting of an image encoder and separate regression heads, failed to produce meaningful predictions, yielding an R^2^ of approximately 0 (MSE 6.1). In contrast, the language-only model achieved a respectable R^2^ of 0.51 (MSE 2.9), indicating that the TCR data contains significant predictive power. However, the complete AD-LLaVA-3D model substantially outperformed both, confirming that the integration of both modalities is critical for achieving optimal performance. This conclusion was further reinforced on the external OASIS dataset, where the vision-only model again failed (R^2^ ↑ 0, MSE 6.3). While the language-only model showed moderate generalization (R^2^ 0.40, MSE 1.82), the full model’s vastly superior performance (R^2^ 0.82) highlights that the vision modality, when properly fused with textual data, is indispensable for building a robust and generalizable forecasting model.

**Figure 4:**
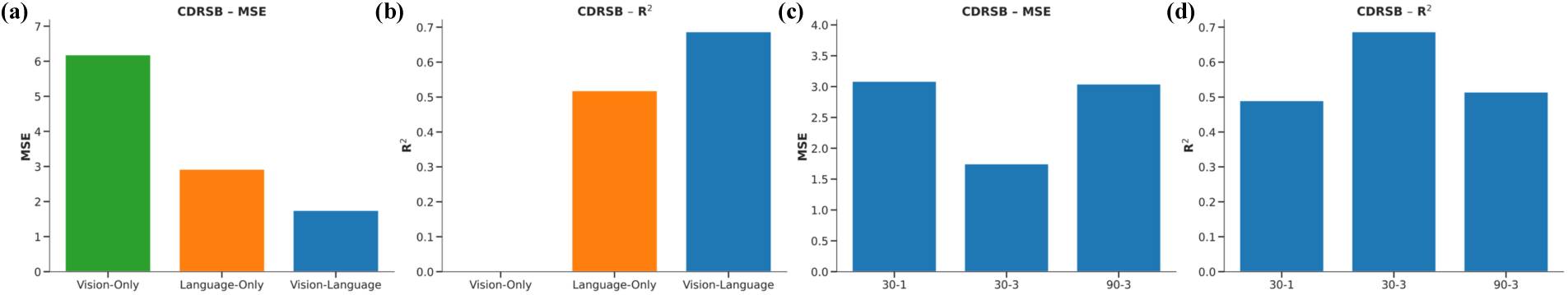
Ablation analysis demonstrating the superior predictive performance of the multimodal integration (Vision-Language) over unimodal baselines, and the impact of different MRI slice selection strategies on forecasting accuracy.

#### 4.4.2 Ablation on MRI Slice Sampling Strategy

For the full multimodal model (MRI + TCR), we further investigated how the amount and distribution of visual information impact performance by testing several slice sampling strategies:

- **Contiguous Slices (30** (**30–1**) **Slices):** Used a dense block of the 30 middle-most axial slices from the MRI scan.
- **Sparse Subsampling (10** (**30–3**) **Slices):** Subsampled every 3rd slice from a central block of 30, resulting in a sparse 10-slice input.
- **Wide-View Subsampling (30 (90–3) Slices):** Subsampled every 3rd slice from a wider central block of 90, resulting in a 30-slice input that covers more of the brain anatomy while maintaining sparsity.

We investigated the impact of MRI slice selection strategy on model performance by comparing three different sampling methods on the ADNI dataset, see Fig 4 (c) and (d). Our primary approach, selecting 10 slices by taking every 3rd slice from the central 30 (denoted as 30-3), yielded the best results with an R^2^ of 0.68 (MSE 2.3). In contrast, denser or wider sampling strategies proved less effective. Using all 30 central slices (30–1) or selecting 30 slices from a wider 90-slice central region (90–3) resulted in significantly lower performance, with R^2^ values of 0.48 and 0.51, respectively. This suggests that the 30-3 strategy strikes an optimal balance between capturing salient information and minimizing data redundancy. Key anatomical indicators for Alzheimer’s disease, such as hippocampal atrophy, are concentrated in specific brain regions captured by the central slices. The superior performance of the 30-3 method implies that providing the model with a downsampled, less correlated set of images from this critical region prevents overfitting to redundant features present in adjacent slices. Furthermore, this sparse sampling approach likely encourages the model to learn more robust and generalizable representations of the 3D neuroanatomical changes, while avoiding the noise introduced by less informative slices included in wider sampling strategies. These combined studies were essential for demonstrating the synergistic benefit of our multimodal approach and for understanding the optimal strategy for processing volumetric medical images.

### 4.5 Implementation and Training Details

All deep learning models were implemented in Python using the PyTorch, huggingface transformers framework and trained on a single NVIDIA A40 GPU with 48GB of VRAM, supported by a system with 128GB of memory and 32 CPU cores. For our proposed AD-LLAVA-3D model, we employed the Low-Rank Adaptation (LoRA) technique for efficient fine-tuning, testing ranks of 4, 8, 16, 32, and 64, ultimately finding that a rank of 16 yielded the best performance. To process the volumetric MRI data (original shape 256×256×256), we adopted a 30-3 slice sampling strategy to create an information-rich yet computationally manageable input of 10 slices. For our baselines, the ResNet-18-3D model used the same 30-3 input for a fair comparison, while Med-Flamingo, due to its inherent design for 2D images, was provided with 3 representative slices from the central region. The classical regression models (Lasso, RF, GBDT) were implemented using their default settings in Scikit-learn, with missing tabular values imputed with zero.

## 5 Conclusion

This study demonstrates a powerful approach, AD-LLaVA-3D, for predicting AD progression to synergistically analyze MRI scans and TCR data. Our novel prompting strategy effectively translates structured TCR data into a rich clinical context, enabling the model to achieve superior predictive accuracy compared to unimodal baselines. An ablation study confirms the significant value of integrating both anatomical and clinical data. These findings highlight the potential of applying advanced multimodal models to revolutionize disease forecasting, paving the way for more accurate, data-rich predictions for AD and other complex neurodegenerative disorders.

## Data Availability

All data produced in the present study are available upon reasonable request to the authors

## Footnotes

1 This work was done at Genentech.

## References

[1] Wafaa Alakwaa, Mohammad Nassef, and Amr Badr. Lung cancer detection and classification with 3d convolutional neural network (3d-cnn). International Journal of Advanced Computer Science and Applications, 8(8), 2017.

[2] Jean-Baptiste Alayrac et al. Flamingo: A visual language model for few-shot learning. arXiv preprint arXiv:2204.14198, 2022.

[3] Norah Alghamdi, Wennan Chang, Pengtao Dang, Xiaoyu Lu, Changlin Wan, Silpa Gampala, Zhi Huang, Jiashi Wang, Qin Ma, Yong Zang, et al. A graph neural network model to estimate cell-wise metabolic flux using single-cell rna-seq data. Genome research, 31(10):1867–1884, 2021.

[4] Carlos G Ardanaz, María J Ramírez, and Maite Solas. Brain metabolic alterations in alzheimer’s disease. International journal of molecular sciences, 23(7):3785, 2022.

[5] Shruthi Bannur, Stephanie Hyland, Qianchu Liu, Fernando Perez-Garcia, Maximilian Ilse, Daniel C Castro, Benedikt Boecking, Harshita Sharma, Kenza Bouzid, Anja Thieme, et al. Learning to exploit temporal structure for biomedical vision-language processing. In Proceedings of the IEEE/CVF Conference on Computer Vision and Pattern Recognition, pages 15016–15027, 2023.

[6] Chun-Hung Chang, Chieh-Hsin Lin, and Hsien-Yuan Lane. Machine learning and novel biomarkers for the diagnosis of alzheimer’s disease. International journal of molecular sciences, 22(5):2761, 2021.

[7] Qi Chen, Ruoshan Zhao, Sinuo Wang, Vu Minh Hieu Phan, Anton van den Hengel, Johan Verjans, Zhibin Liao, Minh-Son To, Yong Xia, Jian Chen, et al. A survey of medical vision-and-language applications and their techniques. arXiv preprint arXiv:2411.12195, 2024.

[8] Wei-Lin Chiang, Zhuohan Li, Zi Lin, Ying Sheng, Zhanghao Wu, Hao Zhang, Lianmin Zheng, Siyuan Zhuang, Yonghao Zhuang, Joseph E. Gonzalez, Ion Stoica, and Eric P. Xing. Vicuna: An open-source chatbot impressing gpt-4 with 90%* chatgpt quality, March 2023.

[9] Pengtao Dang, Tingbo Guo, Sha Cao, and Chi Zhang. A foundational multi-modal model for few-shot learning. arXiv preprint arXiv:2508.04746, 2025.

[10] Pengtao Dang, Haiqi Zhu, Tingbo Guo, Changlin Wan, Tong Zhao, Paul Salama, Yijie Wang, Sha Cao, and Chi Zhang. Generalized matrix local low rank representation by random projection and submatrix propagation. In Proceedings of the 29th ACM SIGKDD Conference on Knowledge Discovery and Data Mining, pages 390–401, 2023.

[11] Jean-benoit Delbrouck, Khaled Saab, Maya Varma, Sabri Eyuboglu, Pierre Chambon, Jared Dunnmon, Juan Zambrano, Akshay Chaudhari, and Curtis Langlotz. Vilmedic: a framework for research at the intersection of vision and language in medical ai. In Proceedings of the 60th annual meeting of the association for computational linguistics: system demonstrations, pages 23–34, 2022.

[12] Ashkan Ebadi, David J. MacDonald, and Santhosh Muralidharan. Deep learning for multi-modal imaging in alzheimer’s disease. Frontiers in Neuroscience, 15:749–763, 2021.

[13] Yingjie Feng, Xiaoyin Xu, Yueting Zhuang, and Min Zhang. Large language models improve alzheimer’s disease diagnosis using multi-modality data. In 2023 IEEE International Conference on Medical Artificial Intelligence (MedAI), pages 61–66. IEEE, 2023.

[14] Charles K Fisher, Aaron M Smith, and Jonathan R Walsh. Machine learning for comprehensive forecasting of alzheimer’s disease progression. Scientific reports, 9(1):13622, 2019.

[15] Marine Fouquet, Béatrice Desgranges, Brigitte Landeau, Edouard Duchesnay, Florence Mézenge, Vincent De La Sayette, Fausto Viader, Jean-Claude Baron, Francis Eustache, and Gaël Chételat. Longitudinal brain metabolic changes from amnestic mild cognitive impairment to alzheimer’s disease. Brain, 132(8):2058–2067, 2009.

[16] Giovanni B Frisoni, Marina Boccardi, Frederik Barkhof, Kaj Blennow, Stefano Cappa, Konstantinos Chiotis, Jean-Francois Démonet, Valentina Garibotto, Panteleimon Gian-nakopoulos, Anton Gietl, et al. Strategic roadmap for an early diagnosis of alzheimer’s disease based on biomarkers. The Lancet Neurology, 16(8):661–676, 2017.

[17] Zhe Gan, Linjie Li, Chunyuan Li, Lijuan Wang, Zicheng Liu, Jianfeng Gao, et al. Vision-language pre-training: Basics, recent advances, and future trends. Foundations and Trends® in Computer Graphics and Vision, 14(3–4):163–352, 2022.

[18] Markus Hafner, Maria Katsantoni, Tino Kö ster, James Marks, Joyita Mukherjee, Dorothee Staiger, Jernej Ule, and Mihaela Zavolan. Clip and complementary methods. Nature Reviews Methods Primers, 1(1):20, 2021.

[19] Kensho Hara, Hirokatsu Kataoka, and Yutaka Satoh. Can spatiotemporal 3d cnns retrace the history of 2d cnns and imagenet? Proceedings of the IEEE Conference on Computer Vision and Pattern Recognition, pages 6546–6555, 2018.

[20] Iryna Hartsock and Ghulam Rasool. Vision-language models for medical report generation and visual question answering: A review. Frontiers in artificial intelligence, 7:1430984, 2024.

[21] Jui-Chan Huang, Kuo-Min Ko, Ming-Hung Shu, and Bi-Min Hsu. Application and comparison of several machine learning algorithms and their integration models in regression problems. Neural Computing and Applications, 32(10):5461–5469, 2020.

[22] Weichen Huang. Multimodal contrastive learning and tabular attention for automated alzheimer’s disease prediction. In Proceedings of the IEEE/CVF international conference on computer vision, pages 2473–2482, 2023.

[23] Clifford R Jack Jr, Matt A Bernstein, Nick C Fox, Paul Thompson, Gene Alexander, Danielle Harvey, Bret Borowski, Paula J Britson, Jennifer L. Whitwell, Chadwick Ward, et al. The alzheimer’s disease neuroimaging initiative (adni): Mri methods. Journal of Magnetic Resonance Imaging: An Offcial Journal of the International Society for Magnetic Resonance in Medicine, 27(4):685–691, 2008.

[24] Keith A. Johnson, Nick C. Fox, Reisa A. Sperling, and William E. Klunk. Machine learning approaches to alzheimer’s disease: A review. Current Alzheimer Research, 18(1):33–52, 2021.

[25] Bijen Khagi and Goo-Rak Kwon. 3d cnn design for the classification of alzheimer’s disease using brain mri and pet. IEEE Access, 8:217830–217847, 2020.

[26] Diyana Kinaneva, Georgi Hristov, Petko Kyuchukov, Georgi Georgiev, Plamen Zahariev, and Rosen Daskalov. Machine learning algorithms for regression analysis and predictions of numerical data. In 2021 3rd International congress on human-computer interaction, optimization and robotic applications (HORA), pages 1–6. IEEE, 2021.

[27] Byounghwa Lee, Jeong-Uk Bang, Hwa Jeon Song, and Byung Ok Kang. Alzheimer’s disease recognition using graph neural network by leveraging image-text similarity from vision language model. Scientific Reports, 15(1):997, 2025.

[28] Chunyuan Li, Cliff Wong, Sheng Zhang, Naoto Usuyama, Haotian Liu, Jianwei Yang, Tristan Naumann, Hoifung Poon, and Jianfeng Gao. Llava-med: Training a large language-and-vision assistant for biomedicine in one day. Advances in Neural Information Processing Systems, 36:28541–28564, 2023.

[29] Feng Li, Renrui Zhang, Hao Zhang, Yuanhan Zhang, Bo Li, Wei Li, Zejun Ma, and Chunyuan Li. Llava-next-interleave: Tackling multi-image, video, and 3d in large multimodal models. arXiv preprint arXiv:2407.07895, 2024.

[30] Haotian Liu, Chunyuan Li, Yuheng Li, and Yong Jae Lee. Improved baselines with visual instruction tuning, 2023.

[31] Haotian Liu, Chunyuan Li, Qingyang Wu, and Yong Jae Lee. Visual instruction tuning, 2023.

[32] Qing Lyu, Shreya Havaldar, Adam Stein, Li Zhang, Delip Rao, Eric Wong, Marianna Apidianaki, and Chris Callison-Burch. Faithful chain-of-thought reasoning. In The 13th International Joint Conference on Natural Language Processing and the 3rd Conference of the Asia-Pacific Chapter of the Association for Computational Linguistics (IJCNLP-AACL 2023), 2023.

[33] Daniel S Marcus, Anthony F Fotenos, John G Csernansky, John C Morris, and Randy L Buckner. Open access series of imaging studies: longitudinal mri data in nondemented and demented older adults. Journal of cognitive neuroscience, 22(12):2677–2684, 2010.

[34] Colin L Masters, Randall Bateman, Kaj Blennow, Christopher C Rowe, Reisa A Sperling, and Jeffrey L Cummings. Alzheimer’s disease. Nature reviews disease primers, 1(1):1–18, 2015.

[35] Daniel Maturana and Sebastian Scherer. Voxnet: A 3d convolutional neural network for real-time object recognition. In 2015 IEEE/RSJ international conference on intelligent robots and systems (IROS), pages 922–928. Ieee, 2015.

[36] Dastan Maulud and Adnan M Abdulazeez. A review on linear regression comprehensive in machine learning. Journal of applied science and technology trends, 1(2):140–147, 2020.

[37] Jing Miao, Charat Thongprayoon, Supawadee Suppadungsuk, Pajaree Krisanapan, Yeshwanter Radhakrishnan, and Wisit Cheungpasitporn. Chain of thought utilization in large language models and application in nephrology. Medicina, 60(1):148, 2024.

[38] Badiea Abdulkarem Mohammed, Ebrahim Mohammed Senan, Taha H Rassem, Nasrin M Makbol, Adwan Alownie Alanazi, Zeyad Ghaleb Al-Mekhlafi, Tariq S Almurayziq, and Fuad A Ghaleb. Multi-method analysis of medical records and mri images for early diagnosis of dementia and alzheimer’s disease based on deep learning and hybrid methods. Electronics, 10(22):2860, 2021.

[39] Michael Moor, Qian Huang, Shirley Wu, Michihiro Yasunaga, Yash Dalmia, Jure Leskovec, Cyril Zakka, Eduardo Pontes Reis, and Pranav Rajpurkar. Med-flamingo: a multimodal medical few-shot learner. In Machine Learning for Health (ML4H), pages 353–367. PMLR, 2023.

[40] Jiquan Ngiam, Aditya Khosla, Mingyu Kim, Juhan Nam, Honglak Lee, and Andrew Y Ng. Multimodal deep learning. In Proceedings of the 28th international conference on machine learning (ICML-11), pages 689–696, 2011.

[41] Yong-Hao Pua, Hakmook Kang, Julian Thumboo, Ross Allan Clark, Eleanor Shu-Xian Chew, Cheryl Lian-Li Poon, Hwei-Chi Chong, and Seng-Jin Yeo. Machine learning methods are comparable to logistic regression techniques in predicting severe walking limitation following total knee arthroplasty. *Knee Surgery, Sports Traumatology*, Arthroscopy, 28(10):3207–3216, 2020.

[42] Alec Radford, Jong Wook Kim, Chris Hallacy, Aditya Ramesh, Gabriel Goh, Sandhini Agarwal, Girish Sastry, Amanda Askell, Pamela Mishkin, Jack Clark, et al. Learning transferable visual models from natural language supervision. In International Conference on Machine Learning, pages 8748–8763. PMLR, 2021.

[43] Shen Rong and Zhang Bao-Wen. The research of regression model in machine learning field. In MATEC Web of Conferences, volume 176, page 01033. EDP Sciences, 2018.

[44] Philip Scheltens, Bart De Strooper, Miia Kivipelto, Henne Holstege, Gael Chételat, Charlotte E Teunissen, Jeffrey Cummings, and Wiesje M van der Flier. Alzheimer’s disease. The Lancet, 397(10284):1577–1590, 2021.

[45] Boran Sekeroglu, Yoney Kirsal Ever, Kamil Dimililer, and Fadi Al-Turjman. Comparative evaluation and comprehensive analysis of machine learning models for regression problems. Data Intelligence, 4(3):620–652, 2022.

[46] Gary W Small, Linda M Ercoli, Daniel HS Silverman, S-C Huang, Scott Komo, Susan Y Bookheimer, Helen Lavretsky, Karen Miller, Prabha Siddarth, Natalie L Rasgon, et al. Cerebral metabolic and cognitive decline in persons at genetic risk for alzheimer’s disease. Proceedings of the National Academy of Sciences, 97(11):6037–6042, 2000.

[47] Muhammad Tanveer, Bharat Richhariya, Riyaj Uddin Khan, Ashraf Haroon Rashid, Pritee Khanna, Mukesh Prasad, and Chin-Teng Lin. Machine learning techniques for the diagnosis of alzheimer’s disease: A review. *ACM Transactions on Multimedia Computing*, Communications, and Applications (TOMM), 16(1s):1–35, 2020.

[48] Lucas R Trambaiolli, Ana C Lorena, Francisco J Fraga, Paulo AM Kanda, Renato Anghinah, and Ricardo Nitrini. Improving alzheimer’s disease diagnosis with machine learning techniques. Clinical EEG and neuroscience, 42(3):160–165, 2011.

[49] Tingyan Wang, Robin G Qiu, and Ming Yu. Predictive modeling of the progression of alzheimer’s disease with recurrent neural networks. Scientific reports, 8(1):9161, 2018.

[50] Yaoting Wang, Shengqiong Wu, Yuecheng Zhang, Shuicheng Yan, Ziwei Liu, Jiebo Luo, and Hao Fei. Multimodal chain-of-thought reasoning: A comprehensive survey. arXiv preprint arXiv:2503.12605, 2025.

[51] Jason Wei, Xuezhi Wang, Dale Schuurmans, Maarten Bosma, Fei Xia, Ed Chi, Quoc V Le, Denny Zhou, et al. Chain-of-thought prompting elicits reasoning in large language models. Advances in neural information processing systems, 35:24824–24837, 2022.

[52] Jiansong Wen, Elise Thibeau-Sutre, Marcelo Diaz-Melo, et al. Convolutional neural net-works for ad classification with structural mri. IEEE Transactions on Medical Imaging, 39(12):3768–3778, 2020.

[53] Jo-Hsuan Wu, TY Alvin Liu, Wan-Ting Hsu, Jennifer Hui-Chun Ho, and Chien-Chang Lee. Performance and limitation of machine learning algorithms for diabetic retinopathy screening: meta-analysis. Journal of medical Internet research, 23(7):e23863, 2021.

[54] Jingyi Zhang, Jiaxing Huang, Sheng Jin, and Shijian Lu. Vision-language models for vision tasks: A survey. IEEE transactions on pattern analysis and machine intelligence, 46(8):5625–5644, 2024.

[55] Xuan Zhang, Chao Du, Tianyu Pang, Qian Liu, Wei Gao, and Min Lin. Chain of preference optimization: Improving chain-of-thought reasoning in llms. Advances in Neural Information Processing Systems, 37:333–356, 2024.

[56] Zihao Zhao, Yuxiao Liu, Han Wu, Mei Wang, Yonghao Li, Sheng Wang, Lin Teng, Disheng Liu, Zhiming Cui, Qian Wang, et al. Clip in medical imaging: A comprehensive survey. arXiv preprint arXiv:2312.07353, 2023.

[57] Kaiyang Zhou, Jingkang Yang, Chen Change Loy, and Ziwei Liu. Learning to prompt for vision-language models. International Journal of Computer Vision, 130(9):2337–2348, 2022.

