## Supplemental figures and tables for "Forecasting Alzheimer’s Disease Progression with Deep Multimodal Learning: Integration of 3D MRI and Tabular Clinical Records via a Large Vision-Language Model"

### Supplementary Material

#### Supplementary Note 1: Longitudinal Data Characteristics

Real-world clinical datasets are rarely complete. To quantify the irregularity of patient visits, we analyzed the intersection of available time points for both the training (ADNI) and external validation (OASIS) cohorts.

#### A. Internal Training Cohort (ADNI)

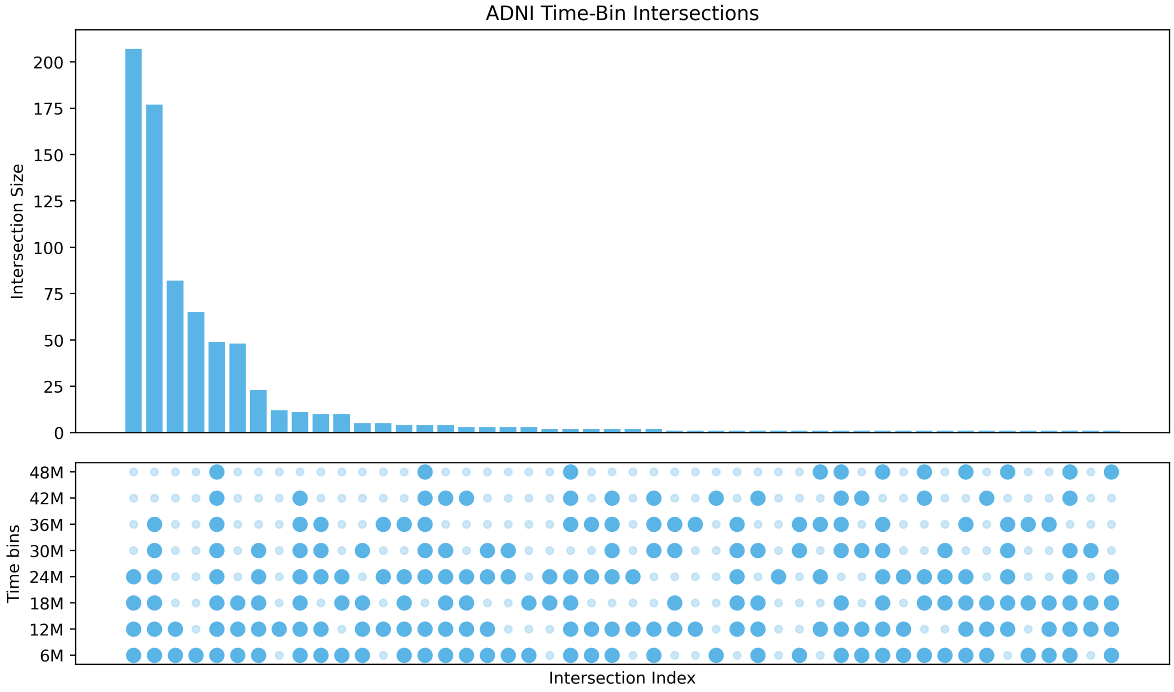


#### B. External Validation Cohort (OASIS)

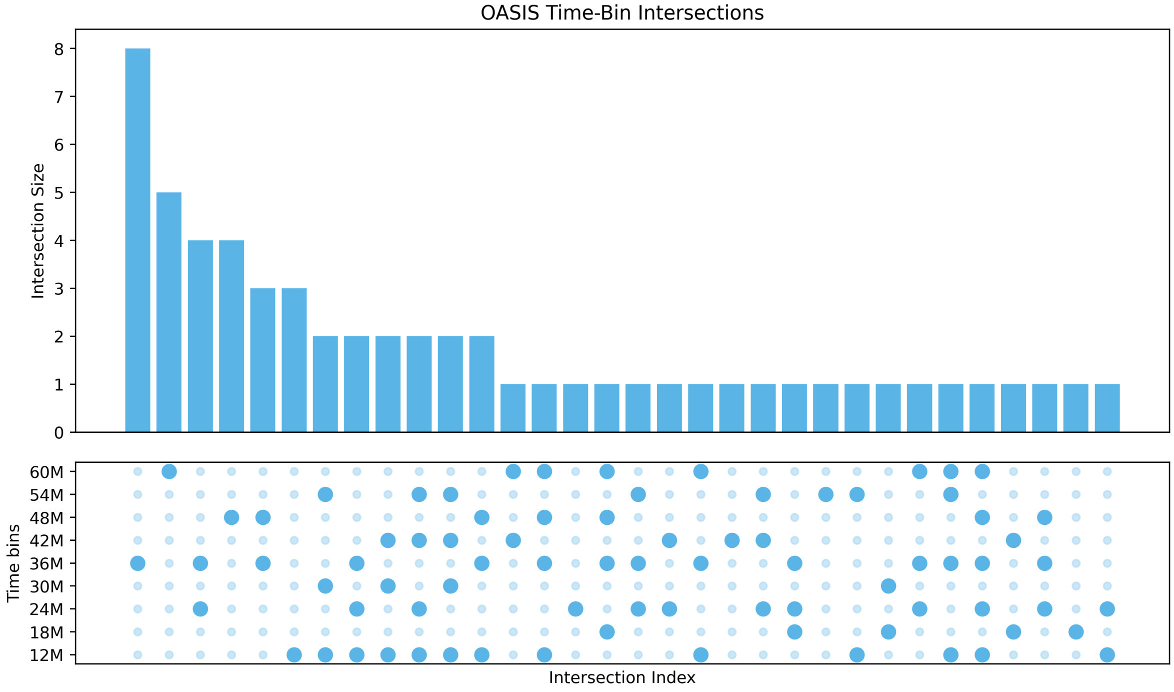




Figure S1: **Visualization of Longitudinal Data Sparsity.** The UpSet plots display the intersection of available visits for each subject. **(Top)** In ADNI, the long tail of unique visit combinations shows high fragmentation. **(Bottom)** OASIS exhibits similar irregularity. AD-LLaVA-3D is designed to

### Supplementary Note 2: Multimodal Prompt Templates

To effectively integrate Tabular Clinical Records (TCR) with volumetric MRI, we utilize a natural language prompting strategy that translates structured clinical variables into a semantic context.

***ROLE AND CONTEXT***

You are a clinical neurologist specializing in neurodegenerative disorders. You are given a patient's current clinical information and their brain MRI scan. Your task is to predict their clinical status after {month diff} months of follow-up.

***CLINICAL CASE PRESENTATION***

Patient {unique random name}, {age}-year-old {gender}, presents with the following clinical information:
Current Status: Diagnosis: {CN/MCI/AD}, CDRSB: {X}, Weight: {XX}, BMI: {XX}, Education Years: {X}, APOE: {X}, MMSE: {X}, Global CDR: {X}

***CLINICAL TASK***

Based on the current clinical presentation and {month diff}-month follow-up period, predict the patient's:
1. Future diagnosis 2. CDRSB score 3. Key cognitive and functional measures

***CHAIN OF THOUGHT ANALYSIS***

Please follow these steps in your analysis:

1. First, analyze the current clinical presentation and identify key risk factors
2. Consider the patient's age progression (from {age i} to {age j} years)
3. Evaluate the impact of age on disease progression
4. Analyze the MRI scan for structural changes
5. Correlate clinical findings with imaging features
6. Consider typical disease progression patterns
7. Formulate your prediction with confidence assessment

***MRI DATA*** <video>

***REQUIRED OUTPUT FORMAT***

Provide your prediction in the following format:
Additional Clinical Information: Global CDR: {X}, BMI: {X}, APOE: {X}, Diagnosis: {CN/MCI/AD}, CDRSB: {X}, MMSE: {X} </s>

Figure S2: Prompt templates 1

***ROLE AND CONTEXT***

You are a comprehensive clinical assessment specialist in neurodegenerative disorders. You are given a patient's complete clinical profile and their brain MRI scan. Your task is to provide a detailed prediction of their clinical status after {month diff} months of follow-up.

***COMPREHENSIVE CASE ANALYSIS***

Patient Information:
ID: {unique random name},
Current Clinical Status: Age: {X}, Gender: {X}, Diagnosis: {X}, CDRSB: {X}, Weight: {X}, BMI: {X}, Education Years: {X}, APOE: {X}, MMSE: {X}

***ANALYSIS OBJECTIVE***

Based on the comprehensive clinical data and {month diff}-month follow-up period, predict: 1. Diagnosis progression 2. CDRSB score 3. Cognitive and functional measures 4. Other relevant clinical indicators

***CHAIN OF THOUGHT ANALYSIS***

Please follow these steps in your analysis:
1. Review complete clinical profile
2. Consider age progression (from {age i} to {age j} years)
3. Evaluate age-related changes and risk factors
4. Analyze comprehensive MRI findings
5. Consider multiple progression scenarios
6. Assess impact of demographic factors
7. Formulate comprehensive prediction

***NEUROIMAGING DATA*** <video>

***REQUIRED OUTPUT FORMAT***

Provide your comprehensive analysis in the following format:
Additional Clinical Information: " Global CDR: {X}, BMI: {X}, APOE: {X}, Diagnosis: {CN/MCI/AD}, CDRSB: {X}, MMSE: {X} </s> "

Figure S3: Prompt templates 2

***ROLE AND CONTEXT***
You are an expert consultant in neurodegenerative disorders. You are given a patient's current clinical data and their brain MRI scan. Your task is to provide an expert opinion on their expected clinical status after {month diff} months of follow-up.

***EXPERT CONSULTATION***
Patient: {unique random name}
Clinical Status: Age: {X}, Gender: {X}, Diagnosis: {X}, CDRSB: {X}, Weight: {X}, BMI: {X}, Education Years: {X}, APOE: {X}, MMSE: {X}

***CONSULTATION REQUEST***
Please provide expert opinion on the patient's expected clinical status after {month diff} months, including: Diagnosis progression, CDRSB score, Cognitive and functional measures

***CHAIN OF THOUGHT ANALYSIS***
Please follow these steps in your analysis:
1. Review current clinical presentation
2. Consider age progression (from {age i} to {age j} years)
3. Evaluate age-related risk factors
4. Analyze MRI findings and their implications
5. Consider disease progression patterns
6. Assess potential interventions
7. Formulate expert prediction

***NEUROIMAGING FINDINGS*** <video>

***REQUIRED OUTPUT FORMAT***
Provide your expert opinion in the following format:
Additional Clinical Information: " Global CDR: {X}, BMI: {X}, APOE: {X}, Diagnosis: {CN/MCI/AD}, CDRSB: {X}, MMSE: {X} </s> "

Figure S4: Prompt templates 3

***ROLE AND CONTEXT***
You are a research scientist specializing in Alzheimer's disease and related disorders. You are given a subject's baseline assessment data and their brain MRI scan. Your task is to predict their clinical progression after {month diff} months of follow-up

***RESEARCH CASE STUDY***
Subject ID: {unique random name}
Demographics: X-year-old female, Baseline Assessment: Age: {X}, Gender: {X}, Diagnosis: {X}, CDRSB: {X}, Weight: {X}, Bmi: {X}, Education Years: {X}, Apoe: {X}, MMSE: {X}, Global CDR: {X}

***RESEARCH OBJECTIVE***
Predict the subject's clinical status after {month diff} months of follow-up, including: Diagnosis progression, CDRSB score, Cognitive and functional measures

***CHAIN OF THOUGHT ANALYSIS***
Please follow these steps in your analysis:
1. Analyze baseline clinical measures and their significance
2. Consider age progression (from {age i} to {age j} years)
3. Evaluate age-related changes in disease progression
4. Analyze MRI biomarkers and their progression
5. Compare with known progression patterns
6. Consider statistical significance of changes
7. Formulate evidence-based prediction

***NEUROIMAGING DATA*** <video>

***REQUIRED OUTPUT FORMAT***
Provide your prediction in the following format:
Additional Clinical Information: " " Global CDR: {X}, BMI: {X}, APOE: {X}, Diagnosis: {CN/MCI/AD}, CDRSB: {X}, MMSE: {X}</s>"

Figure S5: Prompt templates 4

### Supplementary Note 3: Diagnostic Classification Performance

We evaluated the model's capacity to correctly classify patients into diagnostic categories: Cognitively Normal (CN), Mild Cognitive Impairment (MCI), and Alzheimer's Disease (AD).

Table S1: **Diagnostic Confusion Matrices.** Rows represent Predicted classes; Columns represent True classes.

**A. ADNI Test Set (Internal Validation)**

| Pred / True | CN | MCI | AD |
|---|---|---|---|
| CN | 212 | 75 | 2 |
| MCI | 18 | 644 | 125 |
| AD | 0 | 46 | 165 |

**B. OASIS Cohort (External Validation)**

| Pred / True | CN | MCI | AD |
|---|---|---|---|
| CN | 521 | 13 | 1 |
| MCI | 28 | 21 | 10 |
| AD | 0 | 7 | 32 |

### Supplementary Note 4: Extended Cognitive Forecasting

We evaluated forecasting performance on two additional clinical standards: the Mini-Mental State Examination (MMSE) and the Global Clinical Dementia Rating (Global CDR).

### A. Internal Validation (ADNI)

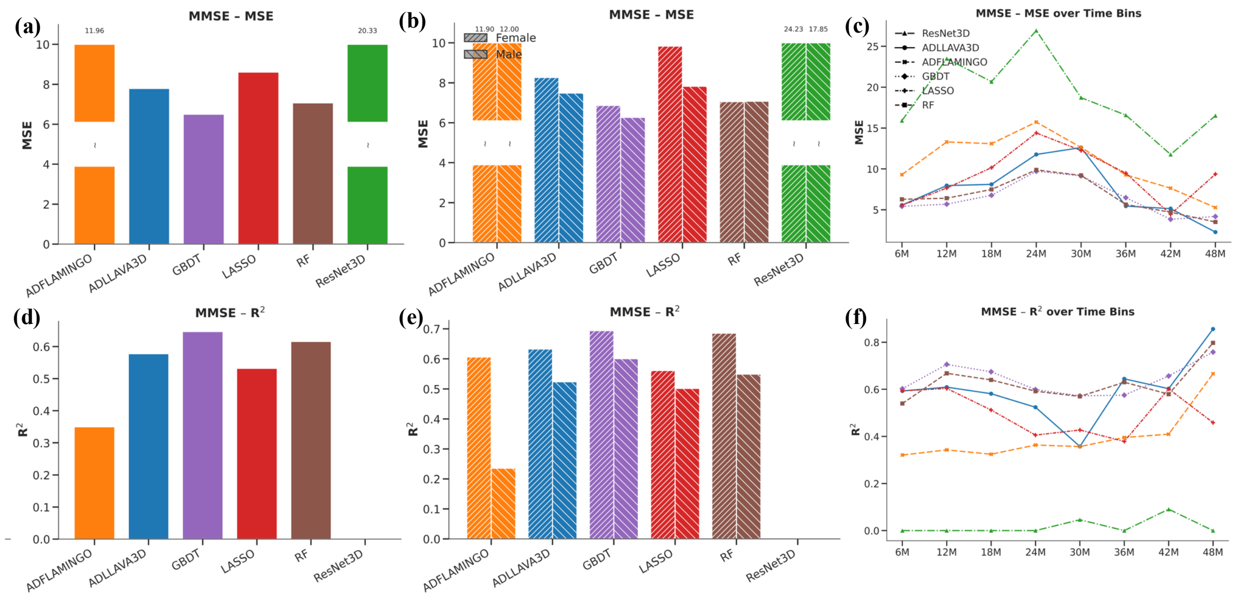


### B. External Validation (OASIS)

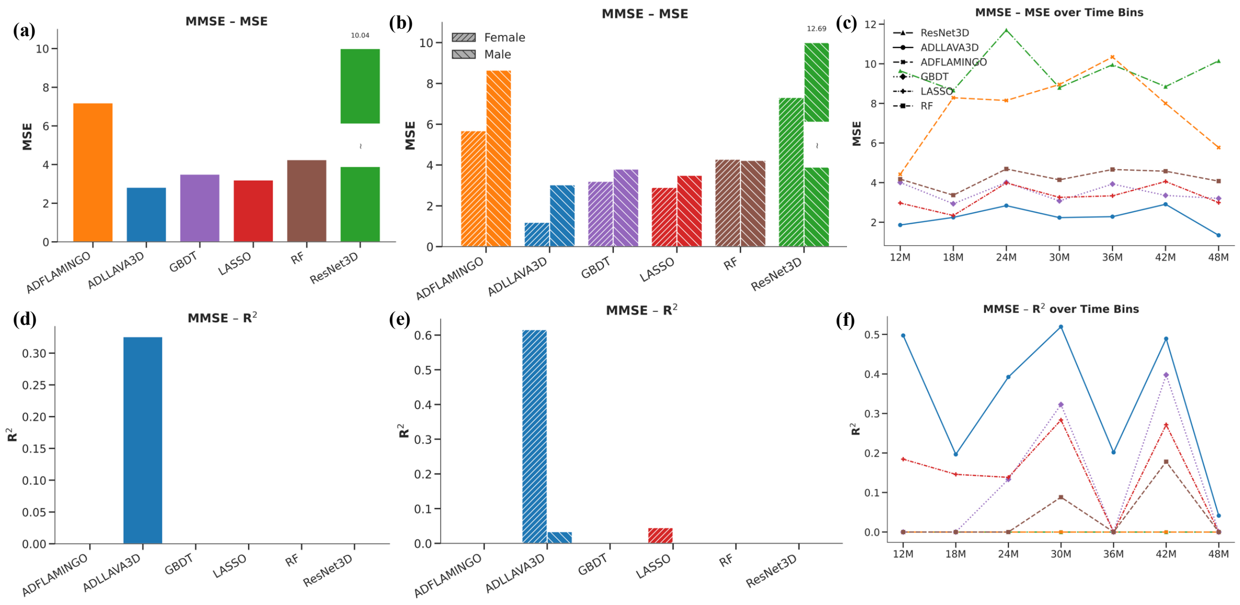


Figure S6: **MMSE Forecasting Performance.** Comparison of predictive accuracy (MSE, $R^2$) and longitudinal stability. Note the "Generalization Gap" in Panel B, where baselines (RF, GBDT) degrade significantly on external data, while AD-LLaVA-3D maintains robust performance.

### A. Internal Validation (ADNI)

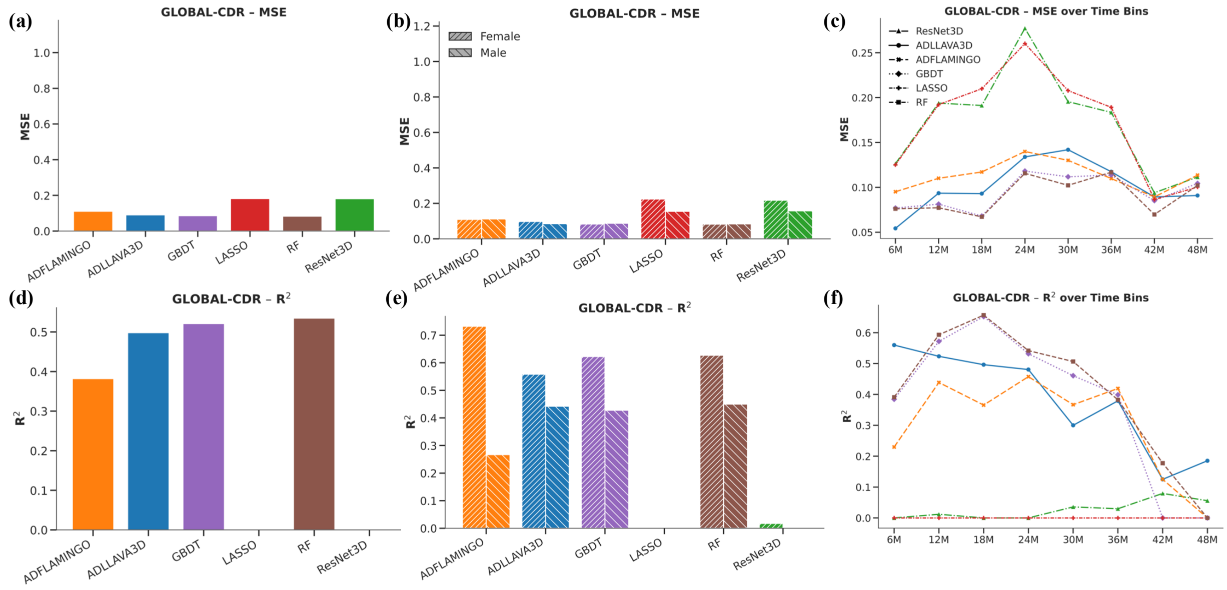


### B. External Validation (OASIS)

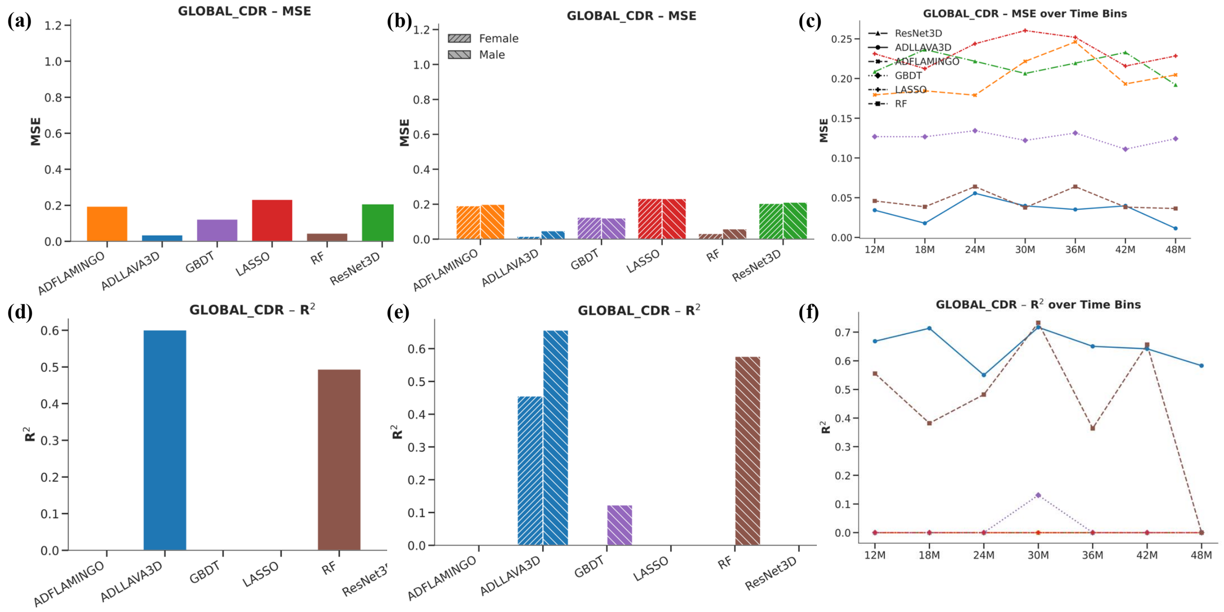


Figure S7: **Global CDR Forecasting Performance.** The proposed model demonstrates superior longitudinal stability compared to unimodal baselines across both cohorts.

### Supplementary Note 5: Biological and Demographic Validation

To verify that the model captures biological signals rather than statistical noise, we tested its ability to predict physiological (BMI) and genetic (APOE $\varepsilon 4$) traits.

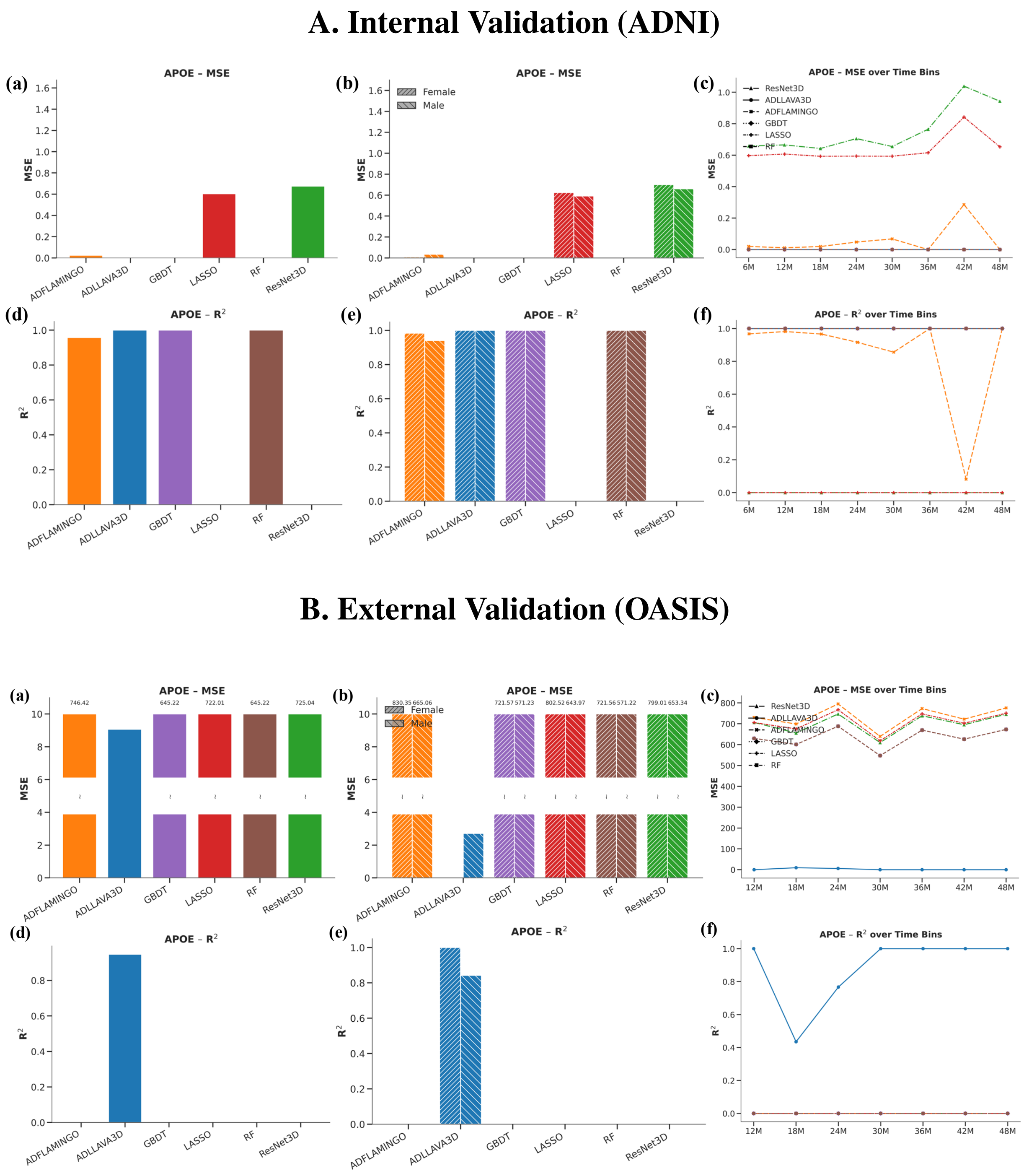


Figure S8: **APOE $\varepsilon$4 Status Prediction.** The high $R^2$ on the external OASIS cohort (Panel B) confirms effective integration of genetic prompts with phenotypic MRI features.

#### A. Internal Validation (ADNI)

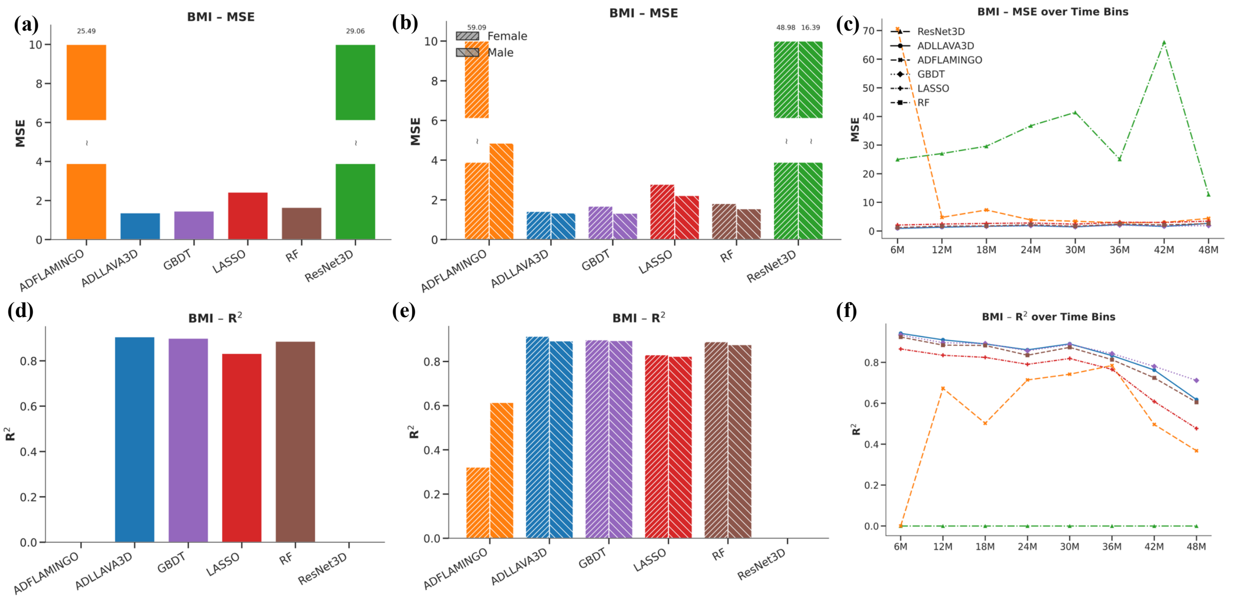


#### B. External Validation (OASIS)

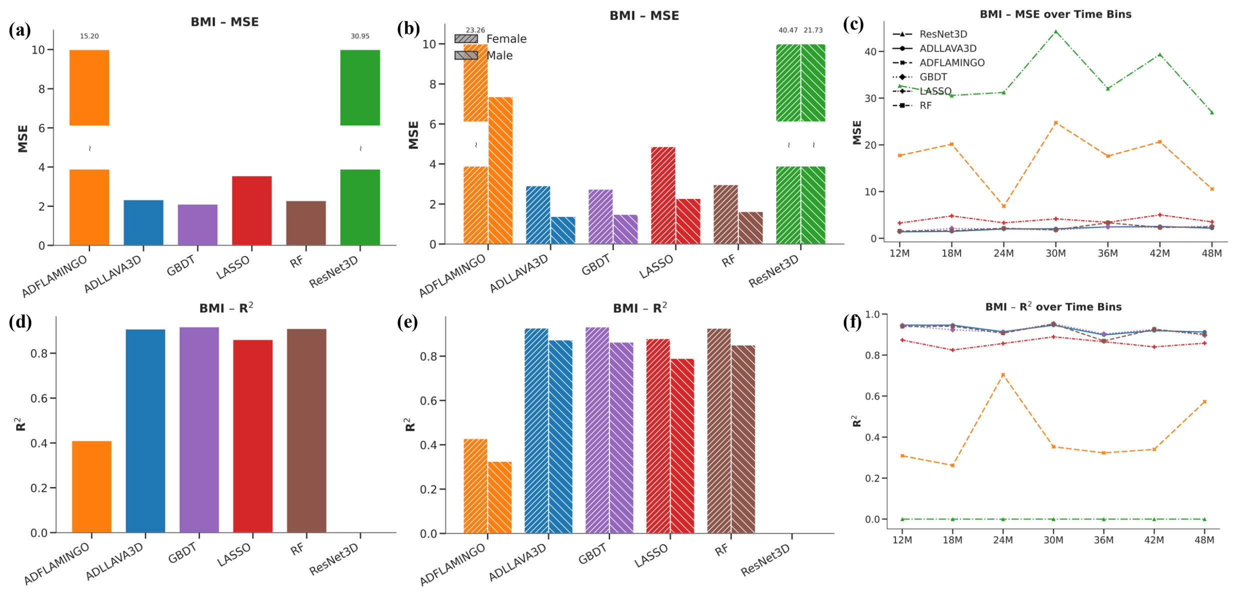


Figure S9: **BMI Regression.** Validation of holistic patient modeling beyond neurological features.

### Supplementary Note 6: Analysis of Generalization Capabilities

A critical finding of this study is the contrast between the ”Generalization Gap” of traditional models and the robustness of ours.

**The Failure of Traditional ”Experts”:** As observed in the external validation panels (Panel B in Figs. S3–S6), traditional models (Lasso, RF) often perform well on ADNI but collapse on OASIS. This indicates overfitting to site-specific distributions.

**The Robustness of AD-LLaVA-3D:** Our framework achieves consistently high performance across both datasets. This is attributed to:

1. **High-Fidelity Inference:** The OASIS validation set underwent strict quality control, allowing the model to perform at its peak potential.

2. **Transferable Features:** By leveraging the semantic reasoning of Large Language Models, the system learns biological concepts of neurodegeneration that remain valid across different hospital systems.
